# MICNet: Prediction of antibiotic susceptibility from microscopic images using transfer learning

**DOI:** 10.1101/2022.04.19.22269518

**Authors:** Adrian Viehweger, Martin Hölzer, Christian Brandt

**Affiliations:** Institute of Medical Microbiology and Virology, University Hospital Leipzig, Leipzig, Germany; Methodology and Research Infrastructure, MF1 Bioinformatics, Robert Koch Institute, Berlin, Germany; Institute for Infectious Diseases and Infection Control, Jena University Hospital, Jena, Germany

**Keywords:** Deep learning, Tranfer learning, Microscopy, Minimum inhibitory concentration, Prediction

## Abstract

Rapid susceptibility testing of bacterial isolates is crucial for anti-infective therapy, especially in critical cases such as bacteriaemia and sepsis. Nevertheless, *empiric therapy* is often initiated immediately and without testing because two days and more pass between a positive blood culture and a susceptibility profile, so in the meantime, the most likely pathogens are treated. However, current empiric recommendations are very generic. They often remain unmodified even in light of incoming, early data specific to a patient’s case, such as positive blood culture microscopy. Part of the hesitancy to change treatments presumably stems from a lack of systematic integration of early information beyond expert intuition. To enable targeted antimicrobial therapy earlier in a case’s progression, we developed a method to predict antimicrobial susceptibility from microscopy images of bacteria alone. Our proof-of-concept `MICNet` combines two neural nets in a new chimerical architecture. It is pre-trained on about 100 thousand antibiograms and fine-tuned with only five thousand microscopic images through transfer learning. Predicting susceptibility profiles of four representative species, we show high predictive performance with a mean F-score of nearly 85%. In addition, several qualitative assessments show that our chimerical net has learned substantial expert knowledge. Therefore, `MICNet` is the first step towards personalized empiric therapy, combining prior pathogen probabilities with patient-specific data.

## Background

In *empiric therapy* treatments are chosen based on the most likely pathogens expected in a particular ailment when the infectious agent is either unknown or incompletely characterized yet. Treatment suggestions are made for patient aggregates, such as “patients with suspected bacteriaemia”, but do not take individual patient characteristics into account. Of course, the clinical microbiologist will modify the treatments suggested in these guidelines on a per-case basis, but this process is often driven by intuition rather than explicit rules. In empiric therapy, treatment is accepted despite the diagnostic uncertainty when a delay in antimicrobial treatment would lead to poor outcomes. For example, in the case of *S. aureus* bacteremia and sepsis, the odds of dying increase by 1.3 % with every hour without treatment.^1^ Usually, there is at least some signal about what agent causes the disease. For example, bacteria can be observed in most cases once a blood culture turns positive. Under the microscope, simple features can be distinguished, such as Gram-stain, shape, and whether the growth was aerobic, anaerobic, or both. These features then lead to inference about plausible pathogens, based on which treatment is initiated.

A human usually performs microscopy without any algorithmic support. However, image analysis has seen an increased use of neural nets in recent years.^2^ They have shown equal or better performance than humans on many tasks,^3^ including in the medical realm, e.g., the classification of skin lesions^4^ and radiographs.^5^ In addition, in microscopy, images of bacterial culture isolates have been assigned a species name with surprising accuracy.^6^ However, such species prediction is only of intermediate interest and arguably could be omitted. We often care about inferred properties of the organism, such as antimicrobial susceptibility, not its name, which only suggests those properties. Recently, MALDI-TOF spectra, now common practice to assign species, have been used to predict antimicrobial susceptibility using a range of algorithms from neural nets and random forests to support vector machines.^7^ Antibiogram prediction from genomes has also been shown in what is known as genotype-phenotype mapping.^8,9^ To our knowledge, microscopic images have not been used to predict bacterial properties, although they are far cheaper to obtain than either spectra or genomes as a diagnostic modality.

We hypothesized that a neural net could extract more information from microscopic images than microbiologists. Subsequently, this information could be used to predict *wild-type* (wt) antimicrobial susceptibility directly. “Wild-type” refers to the expected susceptibility of an organism to different antimicrobials (“antibiogram”), and “susceptible” is defined as a measured minimum-inhibitory concentration (MIC) below some threshold, here so called “breakpoints” defined by the *European Committee on Antimicrobial Susceptibility Testing* (EUCAST, see methods). We do not try to infer resistance at the isolate level, e.g., linking single nucleotide variants to a phenotype. Instead, we aimed to teach a neural net “expert knowledge” from scratch. First, it should recognize different bacterial entities as data-driven latent representations (“embeddings”), not human-defined species. It should do so from a cheap, early diagnostic modality (here images). Then, the neural net should directly link them to expected feature distributions (here MICs). We propose a chimerical model that can do this with high accuracy using transfer learning and that can, in principle, be easily extended to include other diagnostic modalities. Our proof-of-principle opens the way for patient-specific, “personalized” empiric therapy.

## Results

### Unsupervised antibiogram embeddings have plausible global structure

First, we trained a model to “know” about wild-type antibiograms for commonly isolated human pathogens. We had access to an extensive corpus of about 100 thousand antibiograms. For training, we thus selected an unsupervised model, namely a variational autoencoder (VAE).^10^ A significant advantage of the VAE is that it is generative, i.e., once the model has been learned, instances can be simulated from it given a prompt. A VAE represents the input *x* in a constrained latent space (“information bottleneck”) where each instance of *x* has a low-dimensional representation *z* (Figure 1A).

**Figure 1:**
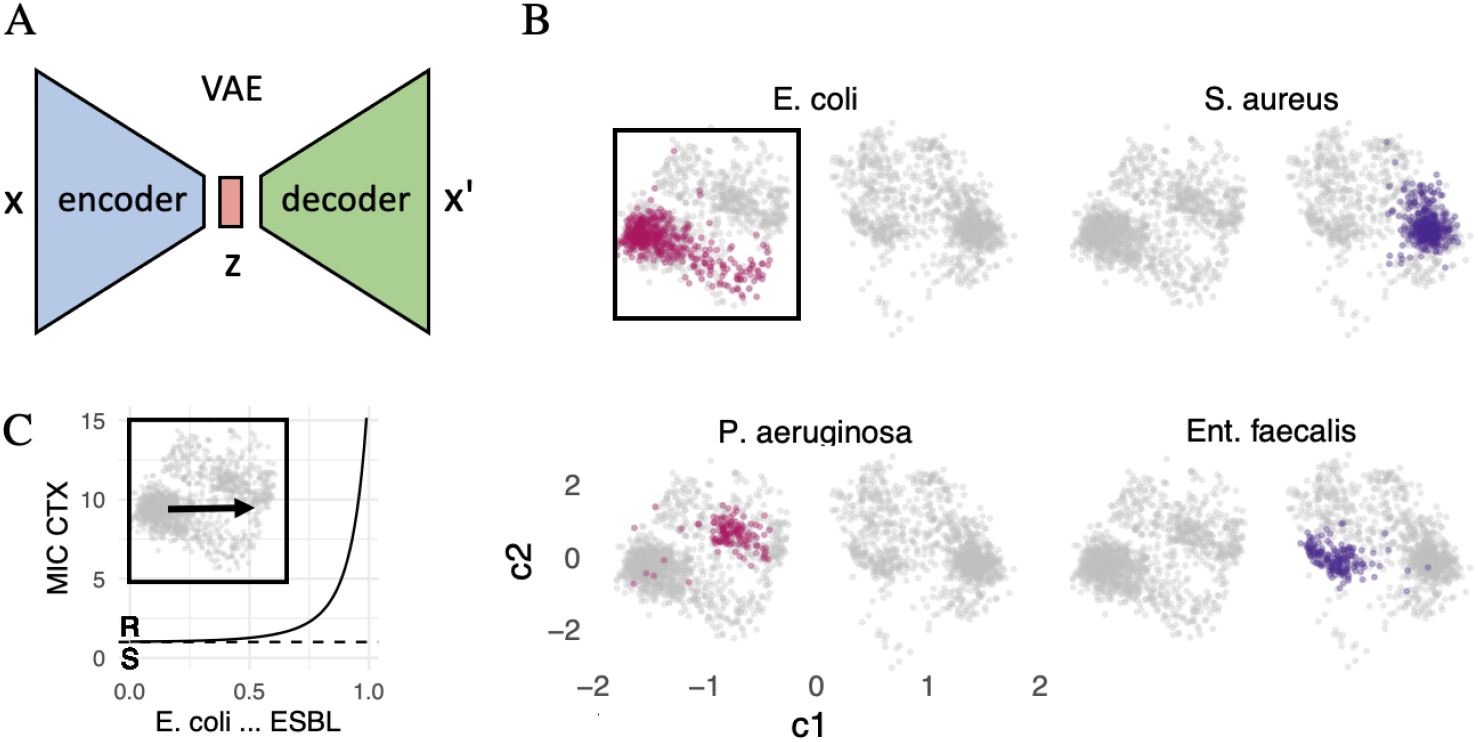
Overview of the two neural nets combined to predict antimicrobial susceptibility from microscopic images. **(A)** A Variational autoencoder (VAE) tries to reconstruct, without supervision, an antibiogram *x′* given the original antibiogram *x* from a low-dimensional “information bottleneck”, a latent representation *z*. After training, the model is generative, and given any value for *z* a MIC *x′* can be reconstructed, independent of the encoder. **(B)** The VAE learns two-dimensional antibiogram embeddings. Displayed are selected Gram-negative (violet) and -positive (blue) bacterial species. Each point is a latent antibiogram representation defined by two coordinates, c1 and c2. Note how, without any species labels, the model nevertheless clusters points by species in latent space. **(C)** In the area of latent space marked in by the black frame in (B), we interpolate along the arrow (insert), using the corresponding two coordinates in latent space *z* to generate an antibiogram *x′* from a point representing an *E. coli* wild-type (wt) isolate, to one that harbors an extended-spectrum beta-lactamase (ESBL). Focusing on cefotaxime (CTX), which ESBL inactivates, we see an exponential increase in its MIC as we move along the arrow. At 1 mg/L an isolate moves from CTX sensitive (S) to resistant (R, see breakpoints for Enterobacterales, EUCAST v11 from 2021). The x-axis displays the interpolation weight *w* of z_ESBL_ between 0 and 1, where 1 *− w* is the weight of wt *E. coli* (z_wt_). The y-axis displays MIC_CTX_ in mg/L.

Training with about 100 thousand MIC profiles (*x*) of 26 antibiotics each resulted in a plausible distribution across latent space. Across all species, antibiogram representations (*z*) of the same species cluster together, similar species are closer than more distant ones (compare *Pr. vulgaris* and *mirabilis*), and Gram-negative and -positive MICs are well separated (Figure 1B and S1). Antibiograms generated from the model (*x′*) are also accurate. For example, in *E. coli*, the wild-type MIC distribution is well approximated (Figure S2). Note, however, how the VAE struggles to model multi-modal or wide distributions because it uses the Gaussian distribution to model data by design (Figure S2).^10^ The variance of the reconstructions is small for similar antibiograms (e.g. *Ent. faecalis* (ATCC 29212), Figure S1). We can assess this using technical replicates in our database (we regularly calibrate the method used to measure MIC, *broth micro-dilution*, using ATCC control strains with corresponding known MIC values).

In the latent space, global distance has meaning, as one interpolates from one point to another (see below). It means that previously unseen values can nevertheless be embedded. We illustrate this in Figure 1C where we record the imputed values for cefotaxim (CTX) moving in latent space from wild-type *E. coli* (MIC_CTX_ *≤* 1 mg/L) to isolates that carry an extended-spectrum beta-lactamase (ESBL), where we expect much higher MIC_CTX_ values. Indeed, we see an exponantial increase in the MIC_CTX_ as we move in the latent space from a sensitive point to a resistant one (arrow in Figure 1C). The same is true for e.g. colistin (COL) when moving from *E. coli* to *Pr. mirabilis* (not shown).

Our qualitative analysis suggests that the VAE learned a plausible representation of antibiograms. However, we did not quantitatively assess the reconstruction loss other than using it as feedback in the training process (i.e., we did not evaluate the VAE on a hold-out set). The reason is that the VAE as an unsupervised algorithm is better evaluated on how well it helps another process, in our case, another neural net. For the latter, validation and test sets were held out according to standard practice (see below).

### Microscopy images can be projected into latent susceptibility space using transfer learning

With the VAE trained, we now had a model which we could prompt with two (latent) coordinates to generate an antibiogram, using only the VAE decoder. Note that the decoder is deterministic, i.e., given a coordinate *z*, it always returns the exact reconstruction *x′*. Next, we sought to project image data into the latent space to generate antibiograms from microscopy images. The inspiration for this architecture comes from the recently published DALL-E model,^11^ which enables zero-shot text-to-image generation.

To test our approach in a proof-of-principle, we used an existing high-quality dataset.^6^ To project images into the (pre-trained) VAE latent space, we used a pre-trained convolutional neural net (CNN) with the ResNet architecture.^12^ The advantage of using two pre-trained models (VAE, CNN), or *transfer learning* more generally, is data efficiency. With them, far less data is required on domain-specific tasks because the models have already learned general features of the data and need only be “fine-tuned” on a few examples to reach good predictive performance.

For all subsequent experiments, we only used a subset of images from four bacterial species, two of them Gram-positive (*S. aureus, Ent. faecalis*) and two Gram-negative (*E. coli, P. aeruginosa*). We chose them because of the clinical relevance to distinguish them in blood culture. For example, Gram-negative rods can be treated empirically with cefotaxime. On the other hand, *P. aeruginosa* cannot be treated with cefotaxime because it is inherently resistant to it. We assigned wild-type antibiograms from our database at random to these subset images. We then fine-tuned the ResNet, which means that we froze all weights but the final layer during training. The final layer (“head”) was replaced with a fully connected layer that outputs two dimensions, compatible with the VAE latent space (Figure 2A). Thus, the neural net has to perform a multi-output regression task. After fine-tuning, the CNN generated embeddings from images that mirrored the VAE. However, a better distinction seems to be possible for the Gram-negative bacteria than for Gram-positive ones (Figure 2B).

**Figure 2:**
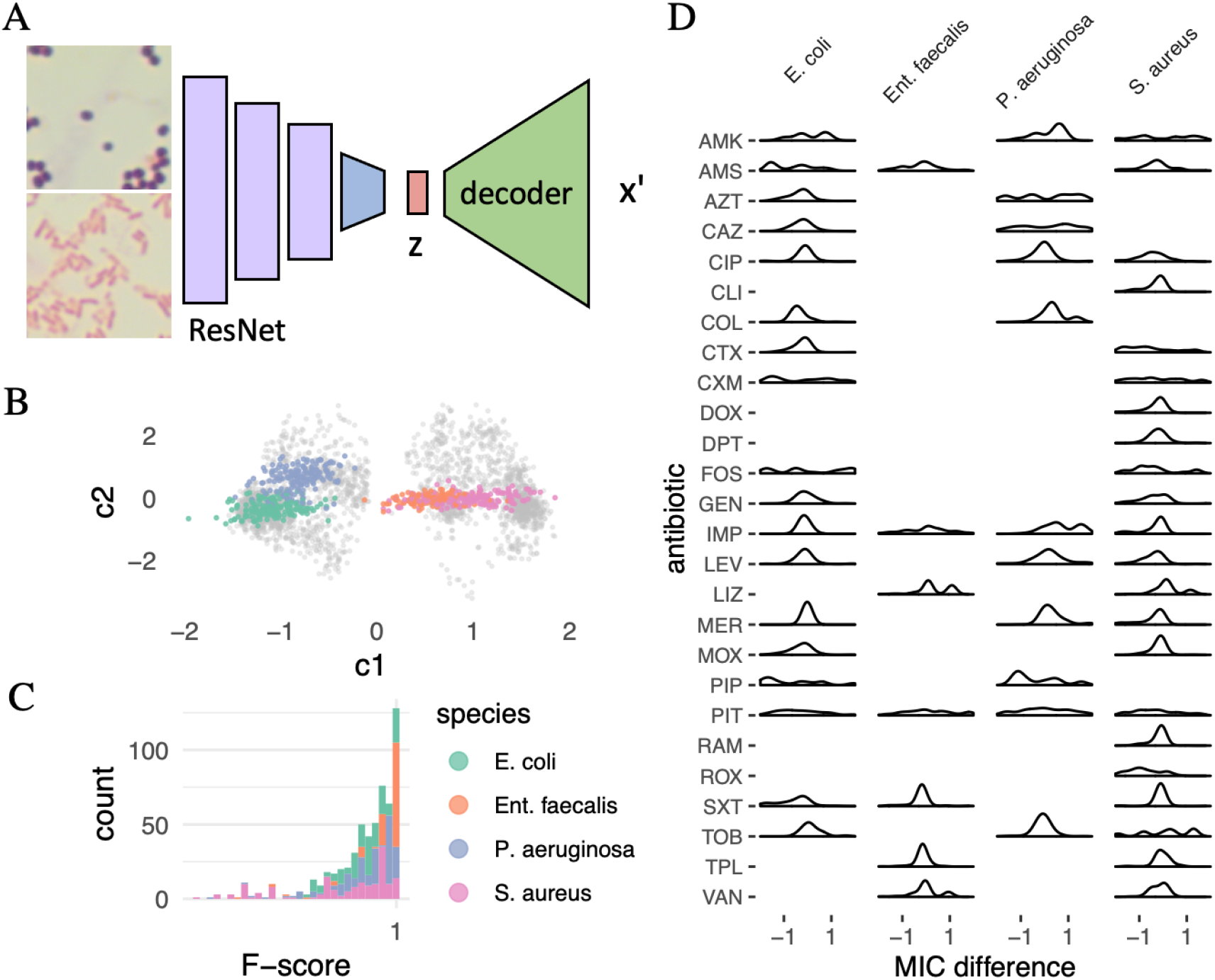
**(A)** Chimerical neural net for transfer learning. A pre-trained convolutional neural net (CNN) is used to encode microscopy images (ResNet architecture, only four of 18 layers are displayed). All layer weights have been frozen during “fine-tuning” (violet) except for the last, fully-connected one (blue). Images are from culture isolates of four bacterial species. Displayed is a Gram-positive photomicrograph in the upper panel and a Gram-negative one in the lower one. The final layer (blue) has been replaced to map the image representation into the latent space of the VAE in Figure 1A (multi-output regression task). The learned image representation *z* is then fed into the pre-trained VAE decoder to generate a MIC reconstruction *x′*. **(B)** Image embedding in the VAE latent space. Each point represents an image from the test set with two coordinates (c1 and c2). *E. coli* and *P. aeruginosa* can be separated well, while *S. aureus* and *E. faecalis* appear more similar to the network. **(C)** Predictive performance of the chimerical neural net in (A) is high. Evaluated to predict the right EUCAST susceptibility class, i.e., whether an isolate was sensitive or resistant, for the entire panel of 26 antibiotics, we achieve a precision of 93.78 *±*12.25 %, a recall of 82.20 *±* 22.32 %, and an F-score of 84.73*±* 17.41 % across all test antibiograms (*n* = 698). **(D)** Distribution of differences of MIC values between original and reconstructed antibiograms. Only antibiotics with corresponding EUCAST breakpoints for the bacterial species were included (EUCAST v11, 2021). For most antibiotics, the difference lies between −1 and 1, which is acceptable to stay within the correct susceptibility class for most substances. Antibiotic abbreviations according to the WHO.

### Chimerical architecture shows good performance in susceptibility prediction task

We evaluated the performance of our proposed `MICNet` architecture in two ways: First, quantitatively, by classifying each predicted MIC value for a given antibiotic and species as correct if it fell in the right breakpoint interval (sensitive, resistant) and calculating precision, recall and the F-score for each predicted MIC profile. Our method reaches a precision of 93.78 *±* 12.25 %, a recall of 82.20 *±* 22.32 %, and a F-score of 84.73 *±* 17.41 % (Figure 2C and Figures S3 and S4). Note that left-skewed distributions can lead to the phenomenon that the sum of mean and standard deviation exceeds the maximum value, here 100 %. The performance is surprisingly good, considering the network can only train on about 3,500 image-susceptibility profile pairs. This frugality illustrates the value of transfer learning whereby the neural net has acquired extensive knowledge about the problem domain through pre-training on data from a different domain (CNN) and unlabelled data (VAE), respectively (see methods).

Second, for a qualitative evaluation, we plotted the distribution of MIC differences for all antibiotics with defined EUCAST breakpoints in the corresponding species (Figure 2D). Most antibiotics fall within the *±*1 range of the true value, which is sufficient to remain in the same susceptibility category (sensitive, resistant). However, on average, the difference is larger for substances with a multimodal MIC distribution in the wild-type, such as fosfomycin (FOS). Furthermore, there are larger differences for antibiotics with a very broad MIC distribution, e.g., piperacillin (PIP, Figure 2D and S2). We address several proposed architectural changes to deal with this shortcoming in the discussion.

## Discussion

We demonstrate how very different modalities can be linked efficiently through neural nets and transfer learning to provide personalized empirical therapy suggestions hours or days before the actual measurement of what was predicted is available.

Interestingly, our proposed chimeric model does not use bacterial species identity or only implicitly through the attached phenotype. We consider this a strength rather than a weakness because the species name is only relevant to predict properties of the organism that are relevant for therapy. Arguably, in clinical microbiology practice, if everything about an isolate was known but the name, not much would change. Also, whether *species* is a useful concept in microbiology remains debated.^13^

Our proposed solution is a proof of principle. We have not scoured the optimization space without prospectively collected data, i.e., blood culture microscopy images combined with MIC profiles. However, we envision several improvements in the following iterations of `MICNet`. First, it can incorporate other data sources available at the same time as microscopy; namely, patient metadata such as age, sex, and current diagnoses^14^ as well as MALDI-TOF spectra of the positive blood cultures.^15^ Such a multimodal net would be trained the same way we did here. Second, we could train the neural net “end-to-end”, propagating the loss from the final generated antibiogram *x*^*I*^ through the VAE decoder and then the CNN. Third, we could optimize both models used. For the VAE, we could use a discretized version, such as the VQ-VAE,^16,17^ as it would model the discrete MIC dilution steps better. For the CNN, we could test other architectures such as a larger ResNet or VGG.^18^ We used microscopy images from bacterial isolate in the current study; with positive blood culture images, image preprocessing will also have to be optimized.^19^

Furthermore, future work will have to include all bacterial and fungal species that can be observed in bacteriaemia. This distribution is highly skewed, which poses a challenge for predicting rare species. However, because our model does not depend on species identification, and the rare species are usually similar to a more abundant one, we assume our model will perform well in this context.

## Conclusions

Our approach proposes a way towards *personalized* empiric therapy, improving clinical outcomes through timely treatment suggestions and minimizing unnecessary prescriptions of antibiotics, which reduces the rate of resistance development.

## Methods

### MIC data aquisition and preprocessing

Susceptibility testing was carried out using the broth micro-dilution method per ISO 20776-1. Broth micro-dilution was performed according to the European Committee on Antimicrobial Susceptibility Testing (EUCAST). Minimum inhibitory concentrations (MICs) for the following antibiotics were determined: ampicillin, ampicillin/sulbactam, piperacillin, piperacillin/tazobactam, ceftazidime, cefotaxime, cefuroxime, aztreonam, imipenem, meropenem, amikacin, gentamicin, tobramycin, ciprofloxacin, levofloxacin, moxifloxacin, colistin, fosfomycin, trimethoprim/ sulfamethoxazole and tigecycline, clindamycin, roxithromycin, vancomycin, teicoplanin, and rifampicin. There are 24 antibiotics in each of two of the routine panels, one for Gram-positive and one for Gram-negative isolates. We merged them into a panel of 26, setting the value for substances not in the panel to 512, i.e., double the highest concentration tested, to signal “resistance” because these substances would not work due to missing targets.

### Image data preprocessing

For micrographs, we relied on the previously published DIBaS dataset,^6^ which was created from pure culture isolates of 33 different genera and species of bacteria for a total of 660 images (equal proportions) at a resolution of 2048 × 1532 pixels (last access 2021-10-01, misztal.edu.pl/software/databases/dibas). To augment the data, we cropped the images into non-overlapping blocks of 224 × 224 pixels, 224 because this is the smallest size compatible with a ResNet architecture (see below). This preprocessing step was also used to avoid excessive blurring due to the CNN pooling layers. Given the original images’ high resolution and “wide-angle”, we would lose important information about shapes and edges. The preprocessed images also mimic how a microscopist would process slides under a microscope, i.e., with smaller fields of view. In line with community standard practice and for ResNet compatibility, the images were loaded in to a range of [0, 1] and normalized in all three RGB channels to means (0.485, 0.456, 0.406) and standard deviations (0.229, 0.224, 0.225) computed from the images originally used to train ImageNet,^20^ and which were subsequently used to pre-train our convolutional neural net (CNN, see below).

### Training, validation, and testing data

From the cropped image data, we selected four species for our experiments. These are relevant pathogens where the distinction is clinically relevant when diagnosing positive blood cultures (see results). From these four species, we randomly selected 70 % (*n* = 3020) of cropped images as training data, and 15 %, respectively, as validation (*n* = 590) and test (*n* = 698) set. We then assigned each cropped image a wild-type MIC of the corresponding species from our database at random. Note how, while this amount of data is hard to collect in practice, it is still an order of magnitude smaller than what would be required to train the models we use from scratch. Only transfer learning using pre-trained weights allows learning from such little data.

### Variational autoencoder model

We trained a variational autoencoder (VAE)^10^ on our entire MIC dataset. Each vector of 26 numbers was forced through a two-dimensional information bottleneck (*z*). We trained for 30 epochs with a mean squared error (MSE) loss, a Kullback-Leibler divergence (KLD) weight of *β* = 1 in line with the original implementation, optimized using Adam^21^ with a learning rate of 0.003 and a batch size of 64.

### Convolutional encoder-decoder model (MICNet) and transfer learning

To leverage transfer learning of images, we used a pre-trained ResNet architecture with 18 layers (downloaded from pytorch vision, v0.10.0, pytorch.org/hub/pytorch_vision_resnet, last access 2021-07-03). We then replaced its final layer (“head”) with a fully connected layer mapping its input to a 2-dimensional vector *ŷ*, i.e., of the same shape as the embedding space of the VAE, and freezing all other weights during subsequent training (“finetuning”). The network thus performs a multi-output regression task. We trained for ten epochs with an MSE loss, optimized using stochastic gradient descent (SGD)^21^ with a (maximum) learning rate of 0.001 and a momentum of 0.9 in a one cycle learning schedule to speed up convergence.^22^ We used a batch size of 64.

For MIC predictions from images, we then fed *ŷ* to the decoder of the VAE to generate MICs deterministically (only the encoder includes a noise term). The final prediction is thus generated from a chimerical network that uses transfer learning to encode and decode the image into a MIC profile. MIC breakpoints used during the evaluation were sourced from EUCAST (v11 from 2021-01-01, eucast.org/clinical_breakpoints). For antibiotics where a breakpoint existed for “susceptible, increased exposure” (I), we used this breakpoint instead of “sensitive”.

## Data Availability

All data produced in the present study are available upon reasonable request to the authors

## List of abbreviations

MIC: minimum inhibitory concentration
CNN: convolutional neural network
VAE: variational autoencoder
NN: neural net
KLD: Kullback-Leibler divergence
SGD: stochastic gradient descent
ESBL: extended-spectrum beta-lactamase
wt: wild-type
EUCAST: European Committee on Antimicrobial Susceptibility Testing

## Availability of data and materials

The MIC data is under restricted access and is not available for public release due to institutional regulations at the University Hospital Leipzig.

## Acknowledgements

We thank Norman Lippmann (Medical Microbiology and Virology, University Hospital Leipzig) for writing the metadata queries our work relies on.

## Supplement

**Figure S1:**
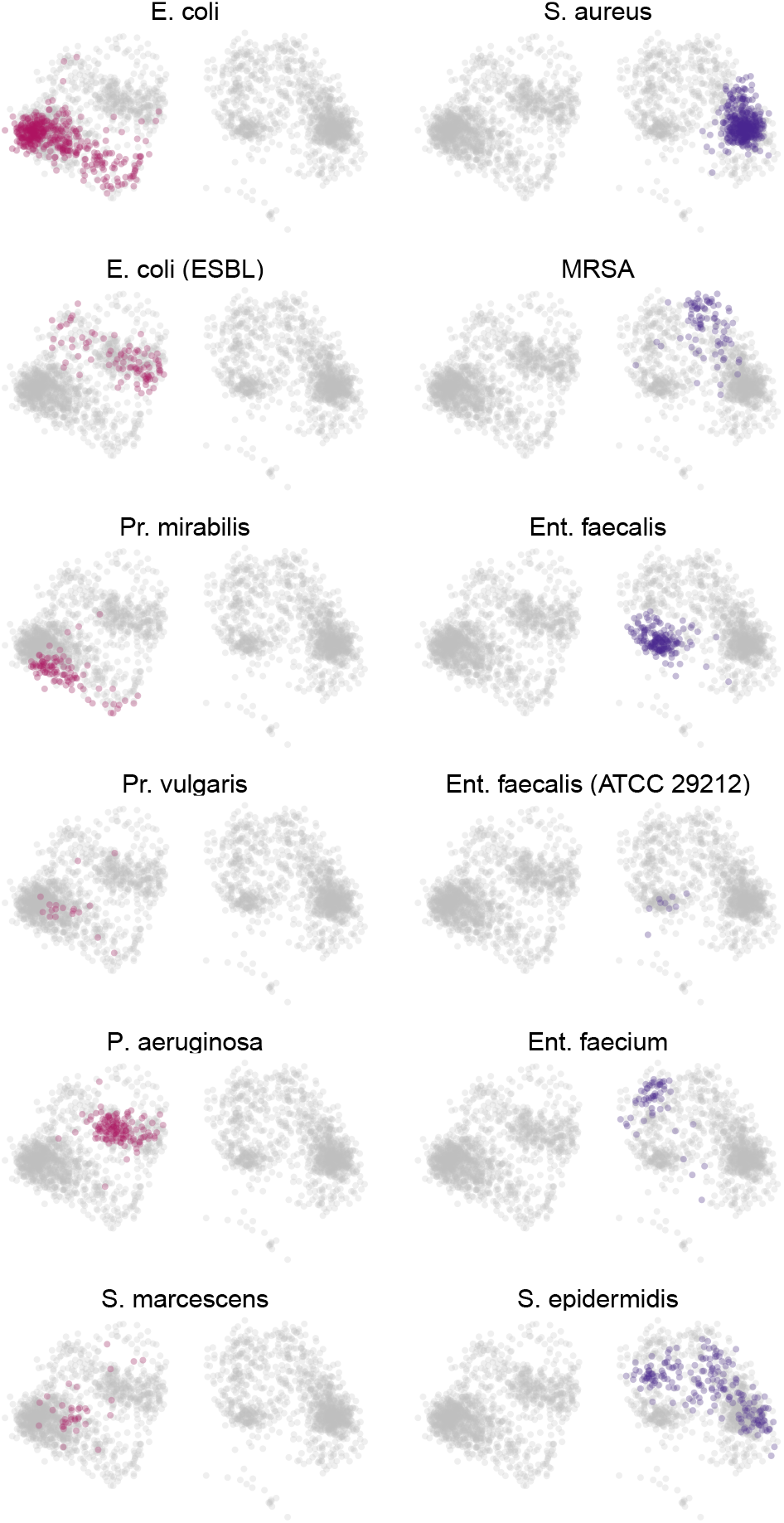
VAE latent space illustrated for additional bacteria.

**Figure S2:**
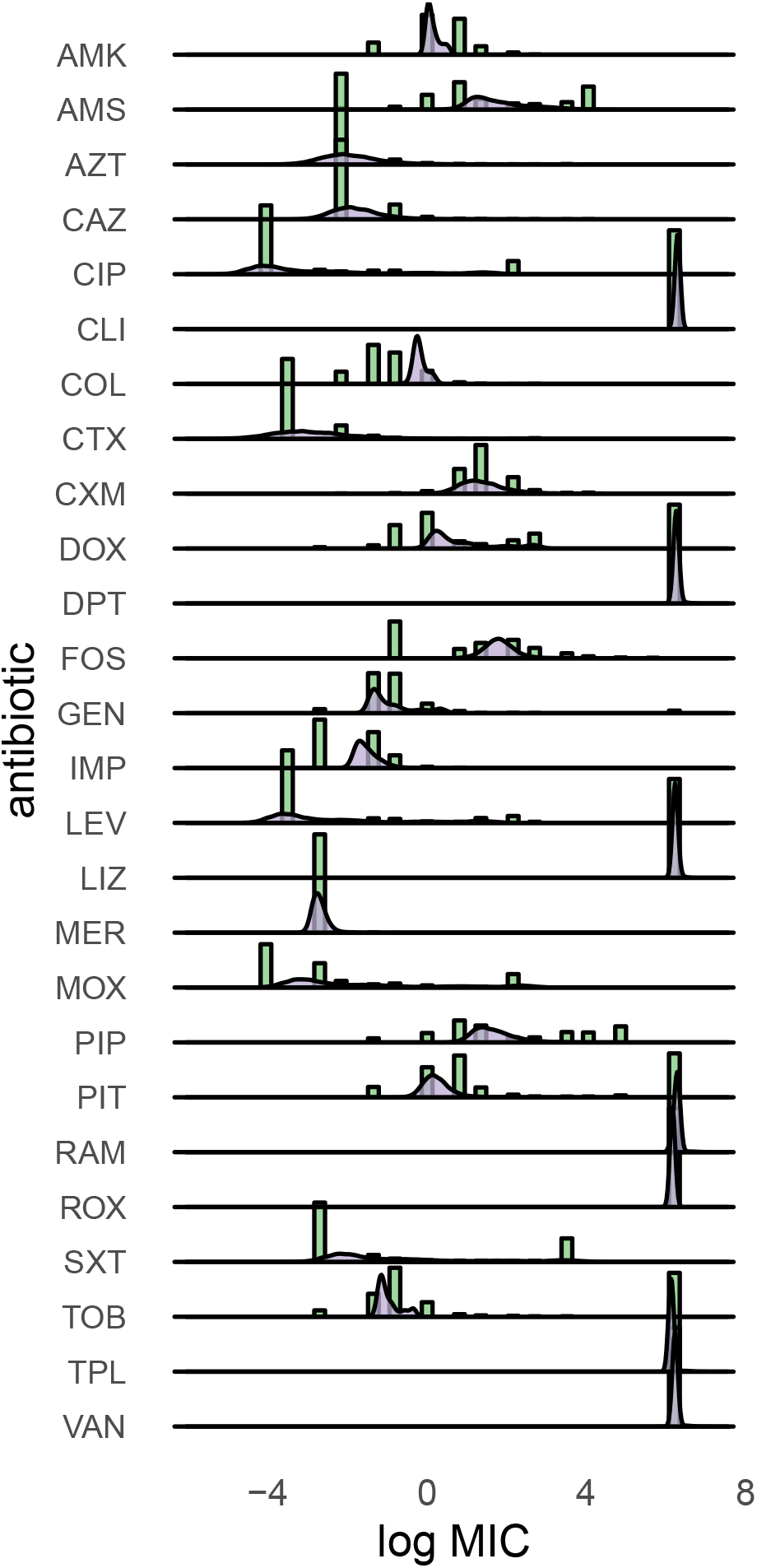
Distribution of MIC values (green histograms) and the distribution learned by the VAE (violet densities).

**Figure S3:**
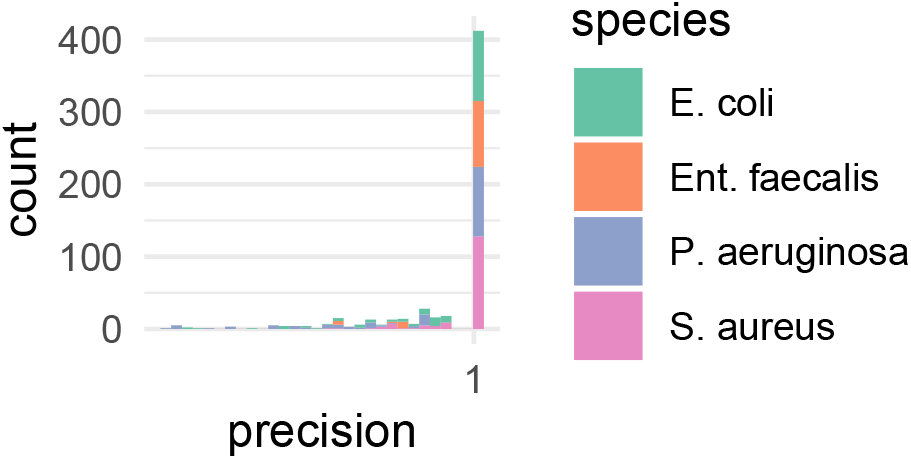
Precision histogram of EUCAST breakpoint class prediction.

**Figure S4:**
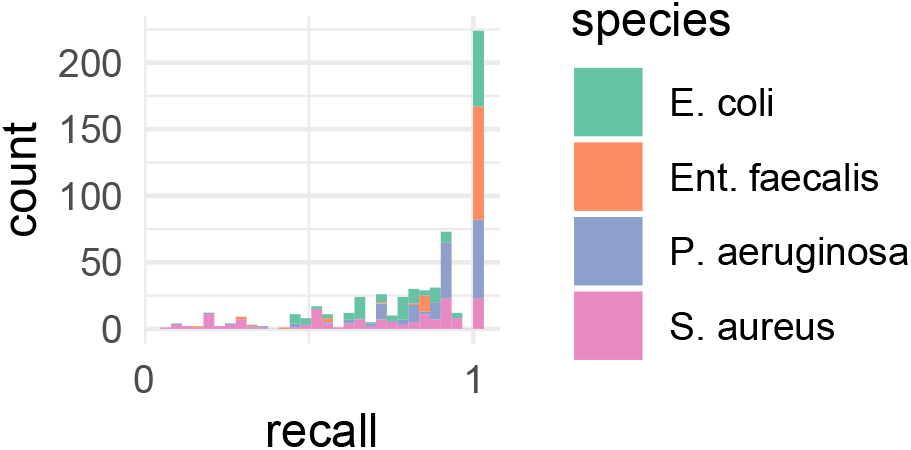
Recall histogram of EUCAST breakpoint class prediction.

